# Association of the rs1042522 SNP with prostate cancer risk: a study of cancer tissues, primary tumor cultures and serum samples from a European Caucasian population

**DOI:** 10.1101/2024.01.07.24300896

**Authors:** Emily Toscano-Guerra, Valentina Maggio, Javier García, Maria Eugenia Semidey, Ana Celma, Juan Morote, Inés de Torres, Marina Giralt, Roser Ferrer, Rosanna Paciucci

## Abstract

Prostate cancer (PCa) is the most common cancer in men and the third leading cause of cancer death in Europe. The *TP53* gene, the most frequently mutated gene in human cancer, is a tumor suppressor gene with crucial functions in preventing tumor development. The single nucleotide polymorphism rs1042522, characterized by the substitution of a proline (PRO) for an arginine (ARG) at the position 72 of the p53 protein (P72R SNP), was studied in 12 primary tumor cultures from prostate biopsies of untreated hormone-naïve patients (hnPCs) with aggressive-metastatic cancer (Gleason ≥8), 11 radical prostatectomies, and a cohort of 94 serums from patient with aggressive prostate cancer using DNA sequencing and melting curve analysis. The results identified a high frequency of the P72R SNP in prostate cancer samples compared to the general European (non-cancer) population, suggesting a very significant association (p<0.0001) between this SNP and the risk of prostate cancer with an odds ratio of 7.937 (IC 95% 5.37-11.00). The G allele (R72) was more frequent in patients with high Gleason scores (≥8) suggesting its association to more undifferentiated-malignant PCa lesions.

## INTRODUCTION

Prostate cancer (PCa), highly prevalent in developed countries, is the most frequent cancer in men and the third cause of cancer death, after lung and colorectal, in Europe^1,2^. In the majority of the cases, the cancer is efficiently eradicated and cured by surgical prostatectomy but late-diagnosed or metastatic aggressive cancers show a five-year relative survival rate below 30%^3^. The tumor suppressor gene *TP53* plays a key role in preventing tumor development. Not surprisingly it has been described as the most commonly mutated gene in human cancers also in PCa^4^. Somatic *TP53* mutations occur more frequently as a later event, and are driver of the aggressive and metastatic cancer, including PCa^5^.

Several hereditary genetic variations, found in approximately 5-20% of different cancer types, are associated with pathogenic variants of genes such *BRCA2* and *ATM*. In PCa, pathogenic germline variants of the *TP53* gene have been described with a relative risk of 4.7-8.6, which is comparable with the frequencies described in those genes^6^.

Germline variants alleles known as a single-nucleotide polymorphisms (SNPs) are present in >1% of the population, occur naturally, and are considered not to have a severe impact on people health. However, many of these variants may impact the structure or the function of the protein.

To study the characteristics of the aggressive metastatic prostate tumors that lead to incurable fatal disease, we established and partially characterized 12 primary tumor cultures derived from prostate biopsies of untreated hormone-naïve patients (hnPCs) with aggressive metastatic cancer. To identify potential predictive markers, the hnPCs cultures were screened for the presence of mutations in driver genes by DNA sequencing. Here, we describe the identification of the *TP53* gene variant allele rs1042522 in the majority of the hnPCs analyzed. The presence of this SNP was confirmed in a cohort of 11 radical prostatectomy tissues and 94 serum samples from Caucasian patients with prostate cancer. The association of the variant allele with prostate cancer risk was investigated using comparisons with the healthy (non-cancer) European (non-Finnish) population from GnomAD v2.1.1.

## MATERIALS AND METHODS

### Prostate primary cultures (hnPCs) from hormone-naïve patients

Primary cultures from prostate tumor needle biopsies of hormone-naïve patients (i.e., without previous treatments) were established in our laboratory from a cohort of patients selected for (**i**) high levels of serum PSA (>50 ng/mL), (**ii**) positive digital rectal examination (DRE), and (**iii**) Gleason ≥ 8. hnPCs were cultured at 37°C in atmosphere of 5% CO2, with complete DMEM-F12 medium, containing: 2 mM of L-Glutamine, 100U of penicillin/mL, 100 µg streptomycin/mL, 0.1 mM of non-essential amino acids, 1 mM of sodium pyruvate and 7% fetal bovine serum, with Supplement 1X, human FGF-10 (10 ng/µL), human EGF (20 ng/µL), and Vitamin A and E (200 ng/µL). Growth factors and vitamins were added freshly whereas supplements were prepared and stored at −20°C until its used. Supplement (100X) was prepared in DMEM-F12 containing glucose (6 mg/mL), transferrin (1000 µg/mL), human insulin (2.500 µg/mL), putrescine (97 µg/mL), sodium selenite (30 µM) and hydrocortisone (100 µM).

### Radical prostatectomies (RPs) samples

Eleven radical prostatectomies (RPs) samples from patients with low-grade operable tumors (Gleason ≤ 7) were used to compare with hnPCs. Samples were collected by the Service of Urology of the Hospital Vall d’Hebron (HVH), preserved in Optimal Cutting Temperature compound (OCT) and stored at – 80 °C until frozen sectioning.

### Serum samples from PCa patients

A cohort of 94 serum samples from patients with diagnosis of prostate cancer were selected with the criteria: PSA values ≥ 50 ng/mL, males southern European. The study was performed with surplus serum samples from routinely tested patients at the Biochemistry Service of HVH, following protocols reviewed and approved by the HVH Institutional Review Board (Medical Research Ethics Committee, protocol number PR(AG) 96/2015).

Serum samples from blood collected in SST yellow tubes containing separator gels, were centrifugated at 3,500 rpm for 15 minutes at 4°C. The resulting serum was collected in 2 mL tubes, properly labeled and codified in the biobank system, were then stored at −80°C. Sample processing was performed according to the scientific and ethical guidelines approved by biomedical research law (Decret 1716/2011).

### DNA extraction

The genomic DNA from hnPCs and RP was extracted using the DNeasy mini kit (Qiagen) following the manufacturer’s guidelines. For the frozen tissue sections from RPs, samples in criotubes were previously mixed with 180 µL of ATL buffer, then homogenized by passing it through a 20-gauge needle attached to an RNase-free syringe about 5-10 times. Successively, samples were incubated with Proteinase K, at 56°C (1 hour) vortexing every 20 minutes during the incubation period. For genomic DNA extraction from serum samples, the MagNA Pure 24 Instrument, and total Isolation Kite (Roche) were used, a fully automated system based on using magnetic glass particles technology (MGP), that allows the processing of up to 24 samples in about 70 minutes.

### DNA sequencing

The presence of the variant rs104252 of the *TP53* gene was first identified in hnPCs and RPs by analysis of the complementary DNA (cDNA) using RNA reverse transcription (RT) and Sanger sequencing. cDNA synthesis was done with RNA (1 µg) previously verified for good quality (RIN ≥ 7), using the NZY M-MuLV kit (NZY Tech) and the 2720 Thermocycler (Applied Biosystems), then the cDNAs were sequenced using Sanger technology.

### DNA genotyping

To analyze the Pro72Arg SNP of *TP53* genotyping was conducted for all samples using the Melting Curve analysis with a customized Light SNiP assay (TIB MOLBIOL) on a LightCycler 480 II instrument (Roche). Reactions were carried out in a final volume of 20 μL, comprising 2.0 μL of LightCycler FastStart DNA Master HybProbe mix (Roche), 1.0 μL of LightSNiP mix, 3 mM MgCl2, and 50 ng of DNA. The ThermoCycler was set up under the conditions outlined in **Table 1**. Subsequently, the melting curves were assessed using the Melt Curve Genotyping software (Roche).

**Table 1.** Cycling conditions for Melting curve analysis.

| Phase. | Cycles | Temperature | Hold | Ramp rate (°C/s) |
| --- | --- | --- | --- | --- |
| Denaturation | 1 | 95 | 10 min | 4.6 |
| Cycling | 45 | 95 | 10 sec | 4.6 |
|  |  | 60 | 10 sec | 2.4 |
|  |  | 72 | 15 sec | 4.6 |
| Melting | 1 | 95 | 30 sec | 4.6 |
|  |  | 40 | 2 min | 2.0 |
|  |  | 75 | 0 sec | - |
| Cooling | 1 | 40 | 30 sec | 2.0 |

### Control group

To study the association of the SNP P72R with prostate cancer, the frequencies found in the (non-cancer) European (non-Finnish) population from GnomAD v2.1.1, containing 134 187 samples, were used as a control group.

### Gleason score and grade groups

The Gleason score is a classification system that assesses the aggressiveness of prostate tumors based on the architecture of the cancerous tissues and the degree of cells differentiation as observed under a microscope. This score, which ranges from 6 to 10, is the sum of two histological patterns (the most common plus the highest grade) within a single tissue sample (**Table 2**). To better understand and describe the cancer aggressiveness, the Gleason score is adjusted into a grade group encompassing values from 1 to 5. The correlation between the score and its respective group, is shown in **Table 2**.

**Table 2.** Prostate cancer classification in groups according to Gleason pattern.

| Patterns* | Score | Group | Significance |
| --- | --- | --- | --- |
| 1+2, 3+2, 3+3 | ≤ 6 | 1 | Low-grade cancer |
| 3+4 | 7 | 2 | Medium-grade cancer |
| 4+3 | 7 | 3 | Medium-grade cancer |
| 3+5, 4+4, 5+3 | 8 | 4 | High-grade cancer |
| 4+5, 5+4 | 9 | 5 | High-grade cancer |
| 5+5 | 10 | 5 | High-grade cancer |

\*Gleason patterns indicate the grade of tissue differentiation.
<sup>1</sup>Small, uniform glands, well differentiated.
<sup>2</sup>More stroma between glands, well differentiated.
<sup>3</sup>Distinctly infiltrative margins, moderately differentiated.
<sup>4</sup>Irregular masses of neoplastic glands, poorly differentiated.
<sup>5</sup>Only occasional gland formation, poorly differentiated/anaplastic tissue.

### Statistics

Analysis of associations of frequencies and proportions were done using Contingency tables and Fisher’s test. Values were considered significant when *p* ≤ 0.05. These analyses were conducted using GraphPad Prism 9 software. Data and image analyses were performed using Circos tool (http://circos.ca/).

## RESULTS

### The *TP53* gene variant allele rs1042522

More than 20 different SNPs have been described in the *TP53* gene^7^, including the variant rs1042522, or P72R, in the amino acid 72 of p53 (**Figure 1A**). This variant contains a guanine at position 357 in codon 72 (C**<u>G</u>**C), a modification that results in a change of the encoded amino acid from proline (C**C**C) to arginine (R or ARG). (**Figure 1B**). **Figure 1C** shows that the arginine induces a decrease in hydrophobicity (turquoise bars) relative to the most abundant proline (P or PRO), which may impact the interaction of the p53 protein with other ligands (CCAR2 or HRMT1L2).

**Figure 1.**
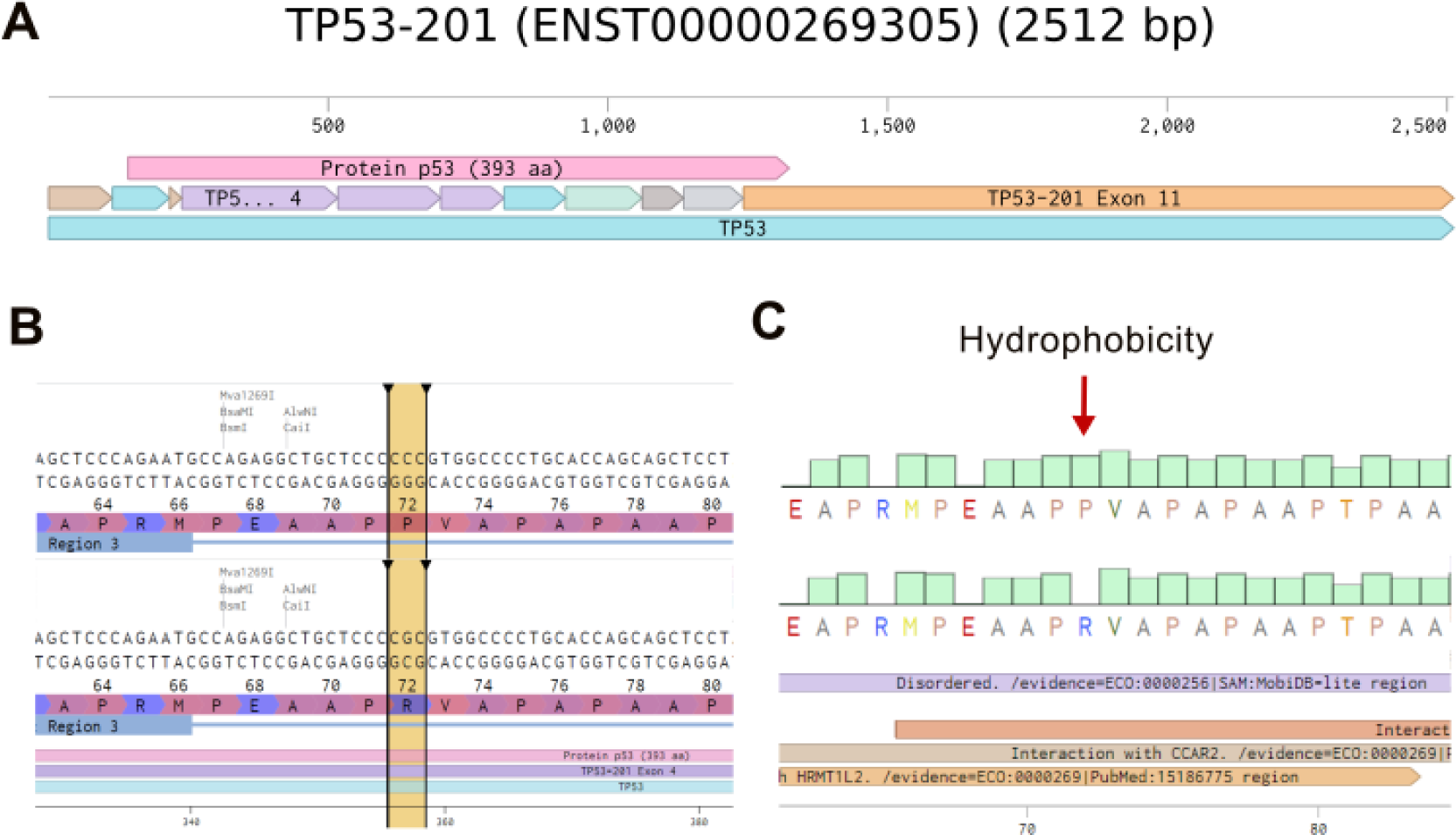
SNP P72R in the TP53 gene. **A)** The canonic transcript for variant rs1042522. The TP53 transcript 201 consists of 11 exons encoding a protein of 393 amino acids including exons 2 to the 5’ region of exon 11. **B)** The SNP P72R (black arrows) polymorphic variants, Proline (P) encoded by the triplet CCC, and Arginine (R) encoded by CGC at position 357 of the coding strand. **C)** The change in the hydrophobic profile of variant rs1042522 (red arrow) when P is changed to R, a polar-positive amino acid. Figures from Benchling.com.

### P72R and allele frequencies in primary prostate tumor cultures and cancer tissues

To study the potential role of *TP53* mutations in the aggressiveness of unresectable prostate tumors, we examined 12 primary tumor cultures (hnPCs) by cDNA sequencing and identified a mutation at position 357 that changed a cytosine to a guanine. This change is not a somatic mutation but corresponds to the allelic variant rs1042522. The frequency found by cDNA sequencing for the ARG variant was 0.82 (9/11) compared to the PRO variant frequency of 0.18 (2/11) (**Table 3**). To confirm these results, the 12 primary cultures plus 11 prostatectomy tissue samples from different prostate cancer patients were genotyped using gDNA melting curve analysis (**Table 3**, **Figure 2A)**. In hnPCs, 2 homozygotes for cytosine (C, proline), 6 homozygotes for guanine (G, arginine), and 4 heterozygotes that predominantly expressed ARG were found. In the RP tissues, 4 homozygotes for cytosine, 5 homozygotes for guanine, and 4 heterozygotes expressed predominantly ARG. The total frequencies of C and G in primary tumor cultures were 0.33 and 0.67, respectively, while in RP tissues the frequency of C was 0.45 and of G 0.55, indicating that primary tumor cultures have a higher incidence of the SNP P72R compared to tumor tissues from prostatectomies. These results would suggest that prostate cancers derived from patients with inoperable tumors may differ from cancers that can be cured by surgery.

**Figure 2:**
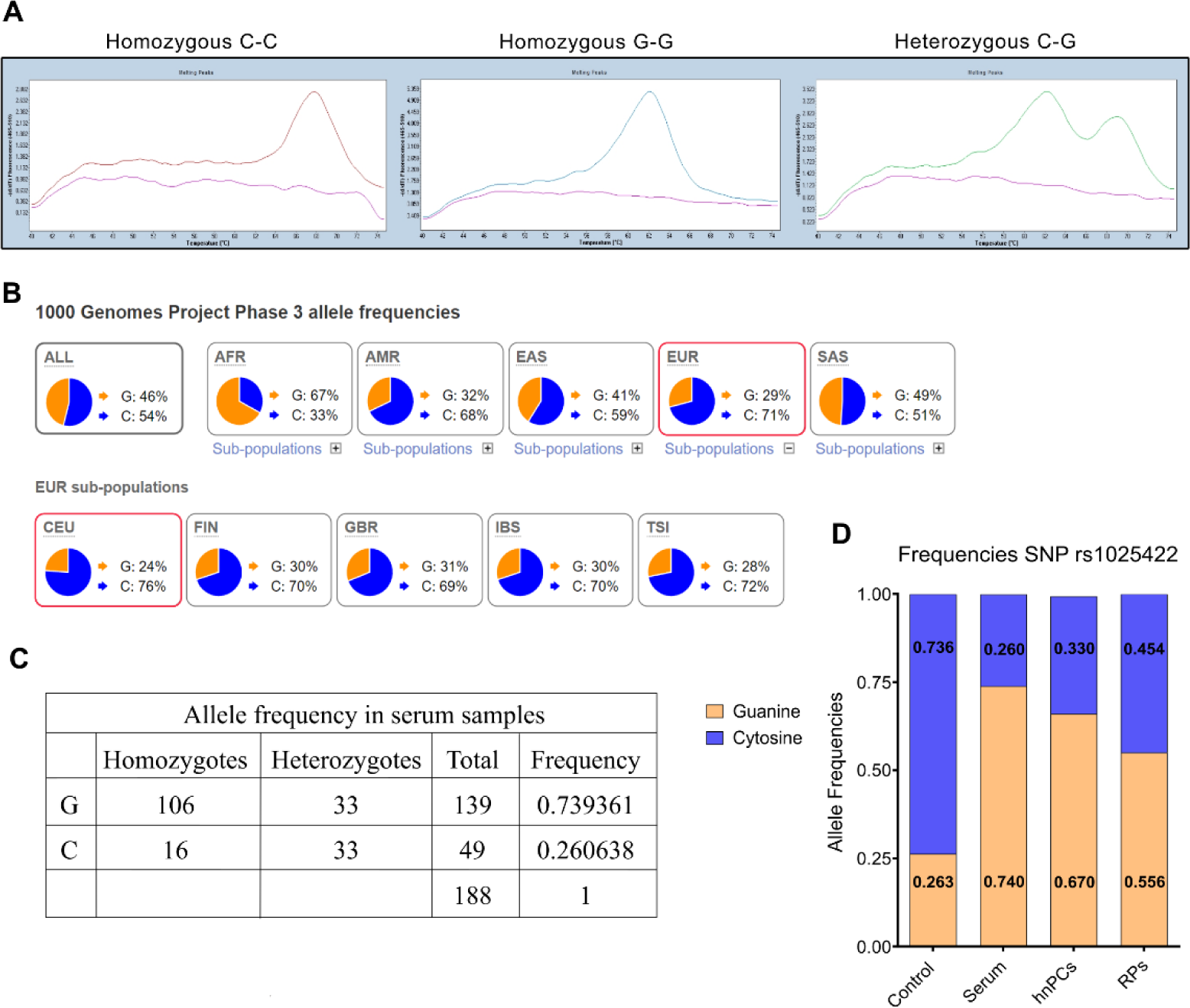
Allele frequencies in the European population and in prostate cancer samples. **A)** Representative melting curves from serum samples. Peaks of absorbance at different dissociation temperatures. Left panel: homozygous samples with cytosine in both strands show a dissociation peak at 68°C. Center panel: homozygous samples with guanine in both strands show a dissociation peak at 60°C. Right panel: heterozygous samples with cytosine and guanine are identified by both peaks. **B)** Global frequency for the allele rs1042522. Allele G has a lower frequency in all sub populations except for the African population. Data and Images from Ensemble.org. **C)** rs1042522 frequencies in serum samples from patients with prostate cancer. **D)** rs1042522 frequencies in primary tumor cultures (hnPCs), serum samples, radical prostatectomy biopsies, compared to the European population.

**Table 3.**
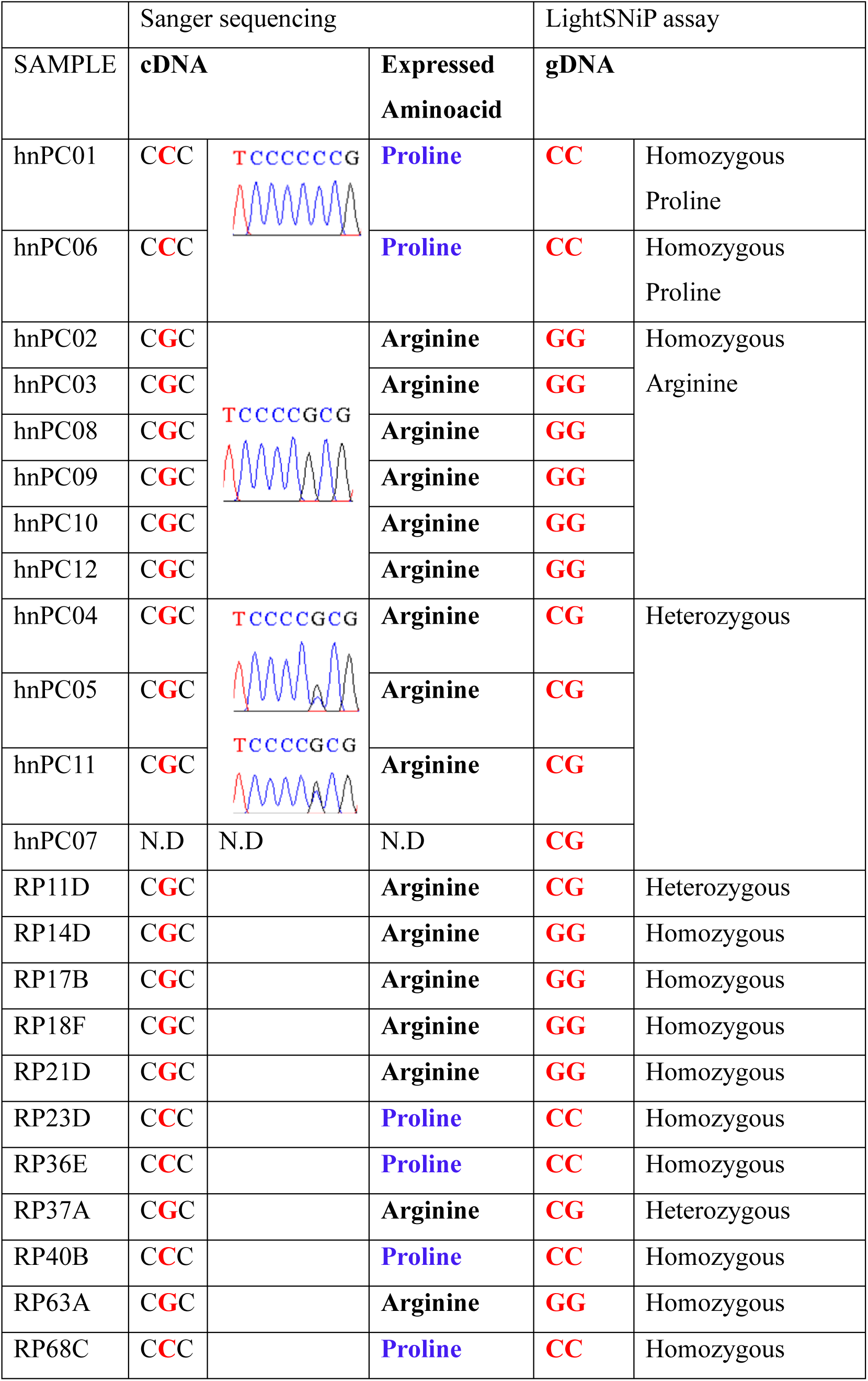
P72R genotyping in primary cultures and radical prostatectomy tissues.

### P72R SNP is associated with prostate cancer risk

To validate the above results, the serum of 94 patients with prostate cancer (PSA ≥ 50 ng/ml) were genotyped (**Figure 2A**). P72R is a controversial SNP, found at different frequencies across populations (worldwide), although the majority of sub-populations show lower frequencies (**Figure 2B**). In the selected European (non-Finnish) non-cancer population the frequency of the G allele was 26.3%, while the frequency of the C allele was 73.6% (**Figure 2B)**. In the serum samples, the frequency of homozygotes for ARG (hARG) was 53.38% (53/94), homozygotes for PRO (hPRO) were 8.58% (8/94) and heterozygotes was 35.10% (33/94) (**Figure 2C).** The allele frequencies observed in our cohort of serum, hnPCs, and prostatectomy tissues were then compared with the (non-cancer) European (non-Finnish) control population (**Figure 2D**). The results show significant differences in the recurrence rate of the P72R SNP in all samples compared to the control population, suggesting an association of the P72R SNP with prostate cancer. The significance of this association assessed by a contingency analysis (Fisheŕs test), showed a very significant association between the P72R SNP and prostate cancer in all samples analyzed, with a frequency of 73.93% for the G allele and an odds ratio of 7.937 (IC 95% 5.37-11.00) for serum samples (p<0.0001) (shown in **Table 4)**.

**Table 4.** Analysis of the association of the rs1042522 variant and prostate cancer.

| Serum samples | Prostate<br>Cancer, n (%) | GnomAD<br>Non-cancer, n (%) | Odds Ratio<br>(95% CI)* | P- value* |
| --- | --- | --- | --- | --- |
| Arginine (G allele) | 139 (0.74) | 31021 (0.26) | 7.937 (5.73– 11.00) | <0.0001 |
| Proline (C allele) | 49 (0.26) | 86797 (0.74) |  |  |
| Total | 188 | 117818 |  |  |
| hnPCs samples |  |  |  |  |
| Arginine (G allele) | 16 (0.67) | 31021 (0.26) | 5.596 (2.395–<br>13.080) | <0.0001 |
| Proline (C allele) | 8 (0.33) | 86797 (0.74) |  |  |
| Total | 24 | 117818 |  |  |
| RPs samples |  |  |  |  |
| Arginine (G allele) | 12(0.55) | 31021 (0.26) | 3.358(1.451-7.772) | 0.0058 |
| Proline (C allele) | 10(0.45) | 86797 (0.74) |  |  |
| Total | 22 | 117818 |  |  |

Contingency table for the association between the P72R SNP and prostate cancer. The selected European (non-Finnish) non-cancer population from GnomAD was used as a control.
\*Odds ratio and p-values were calculated with the Fisher’s Test.

### P72R SNP is not significantly associated with high Gleason tumors

The ability of a mutant p53 protein to neutralize apoptosis and transform cells in cooperation with EJ-Ras is enhanced when codon 72 encodes ARG (R72)^8^. This R72 variant was reported to be able to bind more efficiently and inactivate PGC-1α^4^, a transcriptional target of p53-induced apoptosis in prostate cancer cells, thereby promoting cell invasion and metastasis^4^. These observations support our findings that a higher R72 allele frequency is present in more aggressive PCa tumors. To study whether the Gleason score (grade) of all genotyped patients was associated to higher frequency of the R72 allele, we used the Gleason grading group (1 to 5 instead of 6 to 10) and divided them into 2 different groups: low - medium aggressive tumors (Gleason grade 1 - 3) and highly aggressive tumors (Gleason grade 4 - 5). Patients were genotyped as hARG, hPRO, and heterozygous (ARG-PRO). Data from the cohort of 94 serum patients were plotted on a Circos plot according to Gleason group. **Figure 3A** shows the distribution of patient’s proportions in each Gleason group. Although a marked lower proportion of the P72 allele and a remarkably higher frequency of the R72 allele are visible in high Gleason tumors, these differences did not reach statistical significance (p-value = 0.2867) (**Figure 3A and 3B)**.

**Figure 3.**
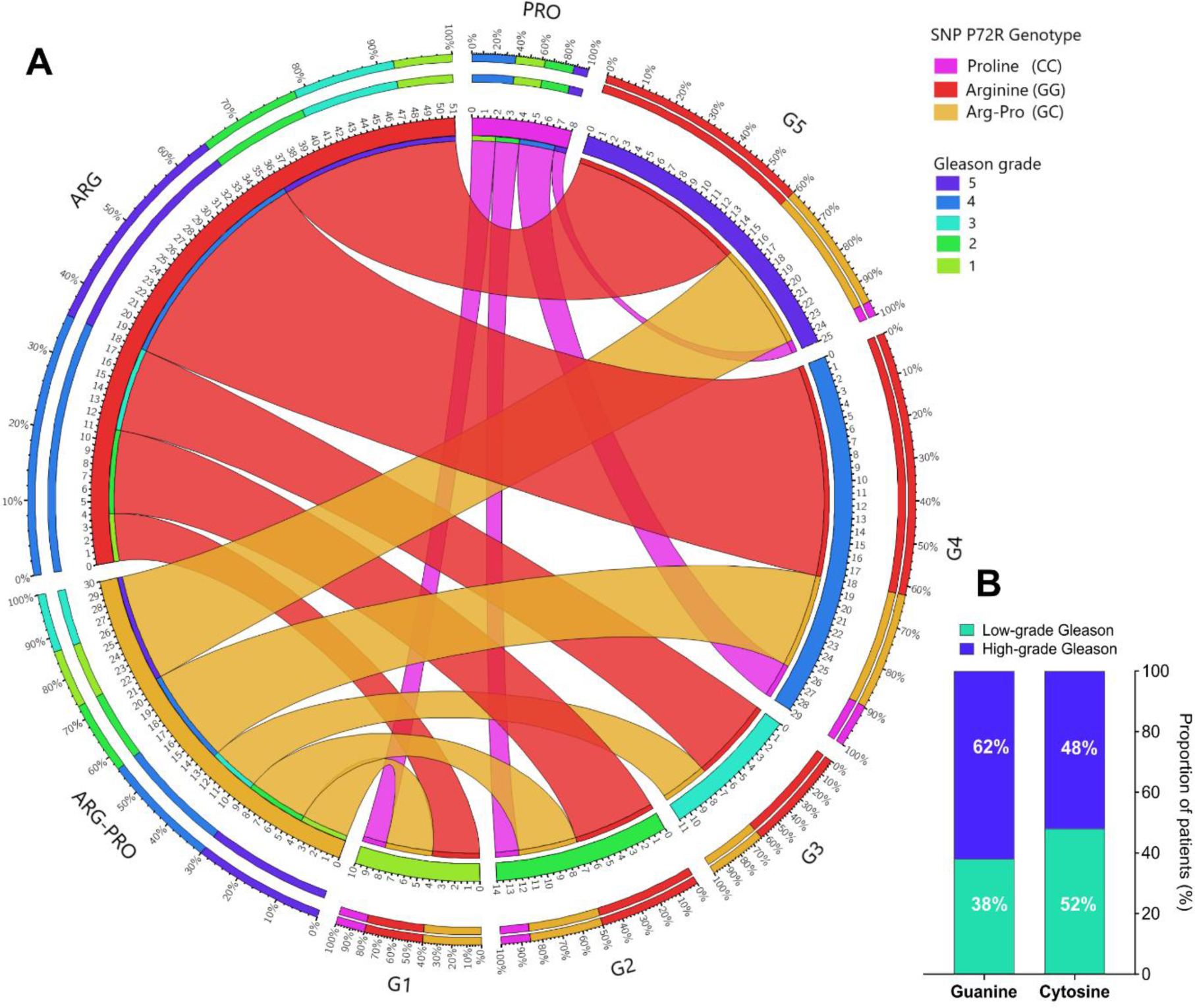
Distribution of the rs1042522 variant in 94 prostate cancer patients according to Gleason grade. **A)** Circos plot displaying the distribution of the P72R genotypes according to patient Gleason score. **B)** Distribution of the SNP in low Gleason grade (1-3) and high Gleason grade (4-5); the contingency analysis of these proportions did not yield significant results. Gleason grade 1: Gleason score ≤6; Gleason grade 2-3: Gleason score = 7; Gleason grade 4: Gleason score = 8; Gleason grade 5: Gleason score

## DISCUSSION

The tumor suppressor gene *TP53,* reported mutated with a high frequency (53.3%) in aggressive mCRPC^9,4^, was the focus of this investigation. The finding that 9 out of 12 primary tumor cultures from patients with aggressive metastatic PCa had a p53 protein with arginine (CGC) at codon 72 while only 2 cultures had a proline (CCC), was intriguing. An initial explanation for the enrichment of the R72 variant in hnPCs from cancer tissues was the Loss of Heterozygosity (LOH), particularly studied in exon 4 that includes the codon 72 of the protein. In fact, loss of the Proline (C) allele and preferential retention of the Arginine (G) allele was described in primary tumors and metastatic tissues of squamous cell carcinoma of vulva^10^, head and neck^11^, colorectal tumors^12^, and lung cancer^13^. In addition, a recent meta-analysis study using the TCGA pan-cancer database and patients selected for the P72R heterozygosity, reported that 31% (127/409) of heterozygotes had lost the P72 allele in the corresponding tumor tissue^14^ indicating that the G allele is preferentially selected for tumor development, possibly due to enhanced p73-induced apoptosis^15^ and the modulation of tumor metabolism by regulating PGC-1α^16^. Although proper LOH screening was not performed in our cancer cohort, the genotyping analysis of serum samples, that showed a similar frequency of the R72 variant compared to hnPCs and cancer tissues from prostatectomy, suggested that it is more likely to be an association with PCa risk.

Because of the high frequency of arginine in the genotyped DNA from hnPCs and RPs was significantly different from the European non-cancer population, a larger number of samples (serum) from patients with prostate cancer were examined. The results confirmed the high frequency of the guanine (C to G) at position 357 compared to a control European non-cancer population selected from the GnomAD v2.1, indicating a very significant association (p<0.0001) of this SNP and prostate cancer risk, with odds ratio of 7.937 (IC 95% 5.37-11.00). Several reports have described a similar significant association of the R72 SNP with the risk of developing cutaneous melanoma, breast cancer, or prostate cancer ^17–20^. The study on PCa compared 187 Iranian patients with 185 control individual negative for cancer^20^. Although the frequency of G allele in cancer patients (65.77%) was not as high as we report here (73.93%), the association was statistically significant compared to the control (p<0.05).

It is worth noting that several reports have shown controversial results^21–23^ or failed to demonstrate a significant association between the rs1042522 variant and the risk of any type of cancer^24–28^, probably due to limited sample size or selection bias. For example, in a colorectal cancer study in the Northern-European population, no significant association between the presence of the SNP and the risk of cancer was reported^29^, but the healthy control samples selected had a high frequency of ARG allele (61%) in contrast to the estimated 26% frequency of the (non-cancer) European population from GnomAD v2.1.1 (**Figure 2**). Similar to our findings, one study in prostate cancer reported an association between Pro/Pro and a lower risk of prostate cancer^30^. However, a number of other reports have shown conflicting results^30–32,27,33–35^. Appropriate representativeness in control samples is necessary for accurate results.

Beyond these conflicting findings regarding the ARG frequency at codon 72 of p53, it is crucial to consider the potential impact of this variant on cancer development, although its full understanding is not clear. Studies in mice revealed that the presence of the R72 variant shows higher incidence of mammary tumors^36^. In addition, this variant associated to increased phosphorylation of p53 and enhanced transactivation of *CDKN1A* (p21^WAF1^) in response to starving, with consequent augmented growth arrest but reduced apoptosis, a situation that favors survival^37^. Furthermore, *TP53* mutations tend to preferentially occur in the R72 allele. In the presence of a mutant *TP53* (*e.g.* R175H, R273H, and A138V) harboring the R72 SNP, an enhanced capacity for migration, invasion, and metastasis in various cancer cell lines, including PCa, has been reported^16^. These observations provide strong evidence that ARG at codon 72 affects the tumor suppression activity of p53.

Although no significant association of P72R SNP with the Gleason score was found in our study, the G allele (R72) was more frequent in patients with high Gleason (≥8, group 4-5) suggesting a possible association to more undifferentiated-malignant PCa lesions (**Figure 3B**). A few studies also evaluated the association between this polymorphism and tumor grades and found no significant association with clinical stage or Gleason grade in PCa^33,34,38,39^. One study identified a link with a modest influence of this SNP and shorter biochemical recurrence (BCR) after radical prostatectomy^35^. Another study in the Japanese population^31^, observed a particularly high frequency of the ARG allele in patients with metastases and those with a high-Gleason score (≥8). However, this study analyzed the association using the arginine allele as a reference, meaning they calculated the significance level between the Pro/Pro and Pro/Arg genotype compared to arginine. As expected, they found no significant difference, as proline did not differ significantly from the control group.

To validate and extend our findings, further investigation with a larger number of samples and consideration of the family history of prostate cancer would be needed. Nevertheless, our present results suggest that a simple but powerful test (gDNA assessment for the P72R SNP) may be useful for the early identification of patients at risk of aggressive metastatic prostate cancer, allowing clinicians to intervene at an early stage.

## Data Availability

All data produced in the present study are available upon reasonable request to the authors

## Acknowledgments

The authors extend their gratitude to the Biochemistry service from Hospital Vall d’Hebron, Barcelona-Spain, for technical support.

## Author contributions

Contributors ETG, and RP conceived and designed the study. ETG, VM, MES, AC and MG collected clinical samples and data. ETG, VM and JG, performed experiments, and analyzed data. ETG and RP generated figures and tables and wrote the manuscript. RP, JM, IT and RF contributed to data interpretation and critically reviewed the manuscript. All authors approved the final manuscript for submission.

**The authors declare no conflicts of interest.**

